# Prognosis-related molecular subtypes and immune features associated with hepatocellular carcinoma

**DOI:** 10.1101/2022.02.13.22270902

**Authors:** Jiazhou Ye, Yan Lin, Xing Gao, Lu Lu, Xi Huang, Shilin Huang, Tao Bai, Guobin Wu, Yongqiang Li, Xiaoling Luo, Rong Liang

**Author notes:** Correspondence to: Rong Liang, Department of Medical Oncology, Guangxi Medical University Cancer Hospital, Nanning, Guangxi 530021, People ’ s Republic of China., Xiaoling Luo, Department of Experimental Research, Guangxi Medical University Cancer Hospital, Nanning, Guangxi 530021, People’s Republic of China., Yongqiang Li, Department of Medical Oncology, Guangxi Medical University Cancer Hospital, Nanning, Guangxi 530021, People ’ s Republic of China. These authors contributed equally to this work (Jiazhou Ye, Yan Lin).

## Abstract

Bioinformatics tools were used to identify prognosis-related molecular subtypes and biomarkers of hepatocellular carcinoma (HCC). Differential expression analysis of four datasets (TCGA, GSE76427, GSE25097, and GSE14520) identified 3,330 genes differentially expressed in the same direction in all four datasets; those genes were involved in the cell cycle, FOXO signaling pathway, as well as complement and coagulation cascades. Based on non-negative matrix decomposition, two molecular subtypes of HCC with different prognosis were identified, with subtype C2 showing better overall survival than subtype C1. Cox regression and Kaplan-Meier analysis showed that 217 of the overlapping DEGs were closely associated with HCC prognosis. The subset of those genes showing an area under the curve > 0.80 was used to construct random survival forest and least absolute shrinkage and selection operator models, which identified seven feature genes (SORBS2, DHRS1, SLC16A2, RCL1, IGFALS, GNA14, and FANCI) that may be involved in HCC occurrence and prognosis. Based on the feature genes, risk score and recurrence models were constructed, while a univariate Cox model identified FANCI as a key gene involved mainly in the cell cycle, DNA replication, and mismatch repair. Further analysis showed that FANCI had two mutation sites and that its gene may undergo methylation. Single-sample gene set enrichment analysis showed that Th2 and T helper cells are significantly upregulated in HCC patients compared to controls. Our results identify FANCI as a potential prognostic biomarker for HCC.

## Introduction

Primary liver cancer is the fourth leading cause of cancer-related deaths worldwide, and it shows remarkable histological and biological heterogeneity [1, 2]. Hepatocellular carcinoma (HCC) is the most common type, accounting for >90% of primary liver cancers [3] and with a 5-year survival rate below 5% [4]. The major risk factors for HCC include infection with hepatitis C or hepatitis B virus, excessive alcohol consumption, smoking, and diet [5]. Genetic risk factors may interact with these environmental risk factors, such as mutations in the genes *CTNNB1* (encoding β-catenin), *TP53*, and *AXINI* [6].

Although early HCC can be treated surgically, recurrence or metastasis may still occur [7]. One major reason is intratumoral heterogeneity, which reduces the efficacy of current therapies aginst other cancers [8]. Sorafenib can significantly improve overall survival (OS) in patients with advanced HCC, but it is extremely expensive and therefore inaccessible to many patients [9]. Identifying genes that help predict HCC prognosis may facilitate personalized treatment and management.

Since HCC is so heterogeneous, tumors in different regions of the body may represent different subtypes [10]. These subtypes can differ in metabolic and signaling pathways, leading to differences in patient survival [11]. For example, one study has identified three HCC subtypes whose tumor microenvironments differ in terms of immune cell compositions [12], and these differences can influence cancer progression [13, 14]. However, only a few studies have identified HCC subtypes based on direct transcriptomic comparison of tumor and non-tumor tissues.

In this manuscript, we used bioinformatics to compare gene expression between tumor and non-tumor tissue in order to identify HCC molecular subtypes and biomarkers to aid prediction of prognosis. In addition, we used single-sample gene set enrichment analysis (ssGSEA) to determine the levels of immune infiltration and characterize the tumor immune microenvironment. Our study identified two molecular subtypes of HCC patients and detected key genes that may serve as potential biomarkers and therapeutic targets for HCC.

## Material and Methods

### Data collection

Gene expression profiles of 371 primary tumor samples, 3 recurrent tumor samples, and 50 normal tissue samples were downloaded from the liver hepatocellular carcinoma (LIHC) dataset in The Cancer Genome Atlas (TCGA) database (https://portal.gdc.cancer.gov/) [15] and from the Gene Expression Omnibus (GEO) database (http://www.ncbi.nlm.nih.gov/geo/) [16]. The GSE14520 dataset included 225 HCC tissues and 220 non-tumor tissues and was obtained based on the GPL3921 platform. The GSE76427 dataset included 52 adjacent non-tumor tissues and 115 HCC tissues and was obtained based on the GPL10558 platform. GSE25097 was based on the GPL10687 platform and included 268 HCC tissues and 243 adjacent non-tumor tissues. GSE138178 included 49 HCC tissues and paired adjacent non-tumor tissues and was obtained using the GPL21827 platform, while GSE84006 ontaining 38 primary HCC tissues and paired adjacent non-tumor liver tissues was obtained based on the GPL5175 platform.

DNA methylation data from 47 tumor and 47 non-tumor liver tissues from Peruvian patients were obtained from the GSE136319 dataset based on the GPL13534 platform. The study flowchart is shown in Figure 1.

**Figure 1.**
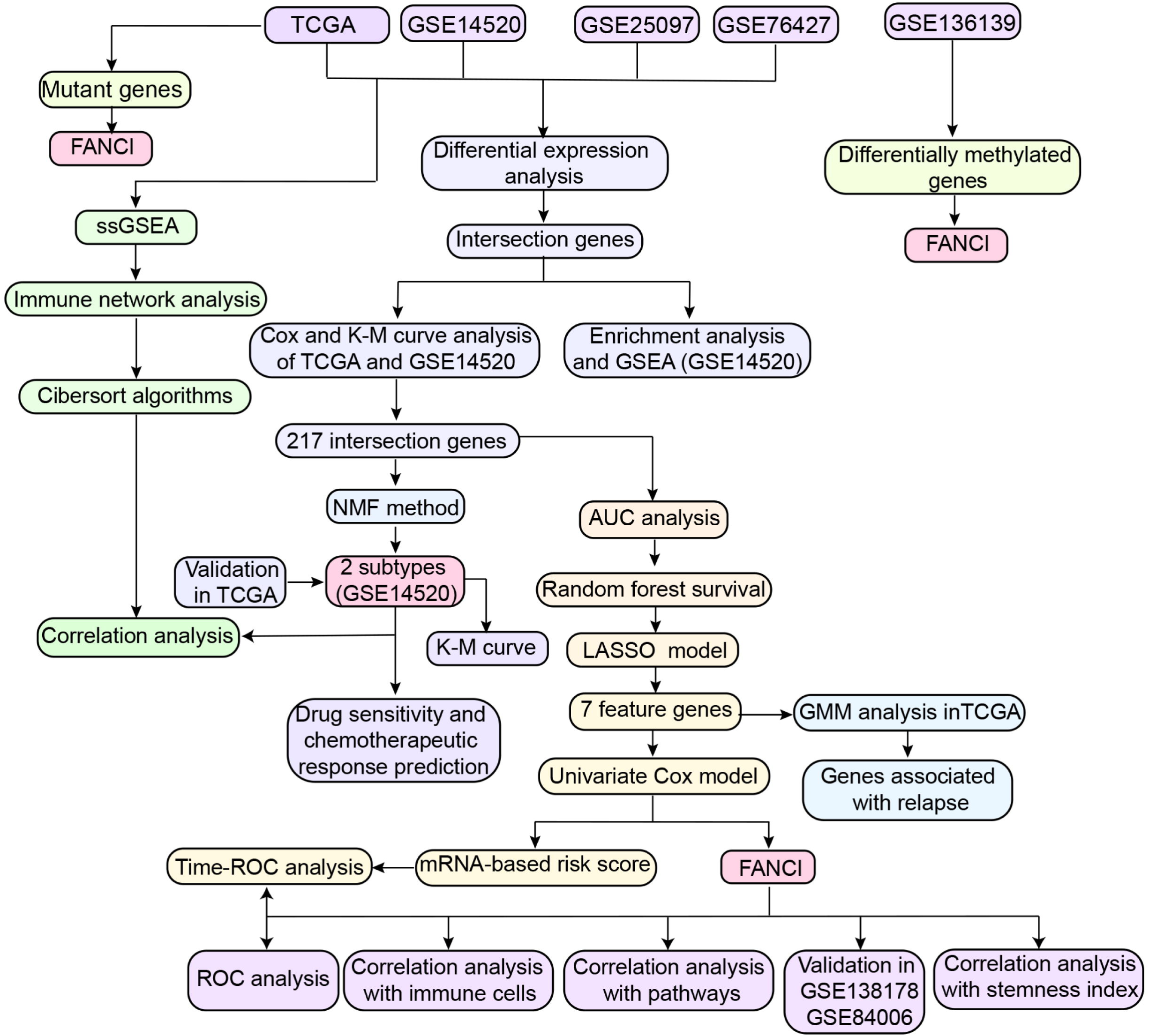
Study workflow. AUC, area under the receiver operating characteristic (ROC) curve; GMM, Gaussian mixture model; KM, Kaplan-Meier; LASSO, least absolute shrinkage and selection operator; NMF, nonnegative matrix factorization; TCGA, The Cancer Genome Atlas; ssGSEA, single-sample gene set enrichment analysis.

### Screening for differentially expressed genes (DEGs) and enrichment analysis

The TCGA datasets and GEO datasets (GSE14520, GSE76427, and GSE25097) were screened for differentially expressed genes (DEGs) between HCC and normal tissues using the *limma* package in R [17]. Statistically significant DEGs (*P* < 0.05) were then identified, and DEGs differentially expressed in the same direction across the four datasets were analyzed for enrichment of Gene Ontology (GO) functions (included molecular function, MF; cellular component, CC; biological process, BP) and Kyoto Encyclopedia of Genes and Genomes (KEGG) pathways using the *clusterProfiler* package in R [18]. Gene set enrichment analysis (GSEA) was then performed, and the results were analyzed using the *fgsea* package in R.

### Cox regression and Kaplan-Meier analyses

Cox regression and Kaplan-Meier curve analyses were performed to identify overlapping DEGs that were significantly associated with OS in the GSE14520 and TCGA datasets. Genes with a hazard ratio >1 or <1 in both datasets based on the Cox regression model were selected, and those genes that also showed a significant effect on survival based on Kaplan-Meier analysis were defined as prognosis-related genes. The Kaplan-Meier curves were used to estimate the conditional survival (CS) of HCC patients, under the assumption that the survival rate for 0-5 years was 100% for patients in the GSE14520 dataset. To determine the prognostic value of the OS-related genes, we used the following formula to calculate the probability of surviving an additional *y* years (CS) given that the patient has already survived for *x* years: CS(x|y) = S(x+y)/S(x), where S(x) is OS at *x* years. For example, the 2-year CS of patients who survived three years after surgery was calculated as CS(3|2) = S(5)/S(3) [19-21]. The functional enrichment of OS-related genes was further verified using metascape (https://metascape.org).

### Construction of random survival forest and least absolute shrinkage and selection operator regression models

A random forest model was constructed based on prognosis-related genes whose area under the receiver operating characteristic curve (AUC) > 0.80 (*P* < 0.01) using the “coxph” function in the *survival* package. To improve the generalizability of the random forest model and reduce overfitting, we reduced the feature dimensions. Features from significant prognostic genes were selected using the *randomForestSRC* package in R, and the genes were classified using the “randomSurvivalForest” algorithm (number of iterations in the Monte Carlo simulation = 100; number of steps forward = 5) [22]. Prognostic genes with a relative importance > 0.2 were considered as final signature genes.

To further reduce model overfitting, we performed least absolute shrinkage and selection operator (LASSO) regression based on the final signature genes using the *glmnet* package [23]. The LASSO regression model is commonly used for high-dimensional prediction [24]. The regression results were processed using the “plotimhistory” function. After 10-fold cross-validation of the parameter selection in the LASSO model, the results were further processed using the “ploidy history” function to obtain feature genes, which were used to calculate the classification efficiency for the 5-year risk score using the *timeROC* package [25].

### Construction of the Gaussian finite mixture model

In order to identify feature genes with strong ability to diagnose HCC recurrence, we constructed a Gaussian mixture model (GMM) using 127 combinations of expression profiles obtained from the TCGA and GSE14520 datasets. The optimal cluster was determined based on the AUC calculated for each model.

### Construction of the feature gene-based risk score prognostic model

Feature genes associated with OS were determined by univariate Cox regression analysis using the *forestpolt* package in R. The risk score for each patient was calculated using the “predict” function of the *survival* package in R [26]. HCC patients were divided into low-risk and high-risk groups based on the median risk score, and their OS was analyzed using the *survival* package. Time-dependent ROC analysis of the GSE14520 and TCGA datasets was performed using the *survival ROC* package in R. Nomograms were plotted using the *rms* package in R, and the consistency index and 95% confidence interval were calculated using the *survcomp* installation package in order to evaluate the predictive power of the model. The results were validated using calibration curves.

### Non-negative matrix factorization

Clustering analysis based on prognosis-related genes was performed using non-negative matrix factorization (NMF) in the *factoextra* package in R (https://CRAN.R-project.org/package=factoextra) and the k-mean clustering algorithm. The average contour width was used to identify the optimal number of clusters. For further validation of the HCC molecular classification, heatmaps were analyzed using the *ComplexHeatmap* and *CancerSubtypes* packages in R [27]. Subtypes were compared in terms of survival analysis.

### Subtype-related drug sensitivity and chemotherapeutic response

To explore the distribution of clinical data (tumor stage, age, sex, and survival time) between the two HCC subtypes in the TCGA and GSE14520 datasets, we used the *dplyr* package in R, and the differences were graphically displayed using the “ggplot2” package in R. A submap algorithm was also used to predict the responsiveness of the HCC subtypes to immunotherapy. If *P* > 0.05, the correlation of the different groups with immunotherapy was considered insignificant. The Tumor Immune Dysfunction and Exclusion (TIDE) database (http://tide.dfci.harvard.edu/) was used to predict the responsiveness of patients to immune checkpoint inhibitors, while the SubMap module of the GenePattern database (https://cloud.genepattern.org/gp) [28] was used to identify similarities between the different subtypes in the GSE14520 and TCGA datasets. *P* values greater than 0.05 indicated high similarity.

The therapeutic response of HCC patients in the GSE14520 and TCGA datasets to anticancer drugs was evaluated based on the Genomics of Drug Sensitivity in Cancer (www.cancerRxgene.org) using the *pRophetic* package in R. The IC_50_ values of the samples were estimated by ridge regression, and 10-fold cross-validation was performed to ensure prediction accuracy. The mean value for duplicate genes was determined using the *allSolidTumors* package in R.

### Gene expression-related stemness index and key gene expression

To calculate the mRNA expression-based stemness index (mRNAsi) in tumor tissues, we constructed a predictive model using the one-class logistic regression algorithm [29]. The mRNA-based signature contained the expression profiles of 10,362 genes. A stemness index model was then used to rank the 211 HCC samples using the Spearman correlation (RNA expression data), and the stemness indices were rescaled to the [0,1] range by subtracting the minimum value and dividing by the maximum value. The HCC samples stratified by the stemness index were used in subsequent integrative analyses. For external validation, the expression of key genes in the GSE138178 and GSE84006 datasets was explored using the Oncomine (https://www.oncomine.org/) [30] and Tumor Immune Estimation Resource (TIMER) (http://timer.cistrome.org/) databases, with selection criteria defined as *P* < 0.001 and fold change > 1.5.

### ssGSEA

The relative levels of immune cell infiltration in HCC and control samples were determined by ssGSEA using the *GSVA* package in R [18]. Correlations among the 24 types of immune cells was then explored by immunity network analysis, while the correlation of feature genes with immune infiltration was assessed by Pearson correlation analysis. The CIBERSORT algorithm was used to quantify the proportions of immune cells in the HCC samples.

### Mutant genes and DNA methylation analysis in HCC

The mutation data of overlapping DEGs in TCGA were visualized and analyzed using the *maftools* package [31], and the position of genetic mutations was determined using the *lollipop* package [32]. Differences in the total number of 450k probes and differentially methylated positions between HCC and control samples in the GSE136319 dataset were also identified. Associations among methylation, gene expression, and clinical phenotypes as well as the correlation between key gene expression and methylation status in TCGA HCC samples were assessed using the MEXPRESS tool (http://mexpress.be/) [33].

## Results

### DEGs in HCC and their functional enrichment

To identify genes related to prognosis in HCC, we first performed differential analysis using data from the TCGA, GSE76427, GSE25097, and GSE14520 datasets (Figures 2A-B). Of the 3,330 DEGs overlapping across the four datasets, 2,058 were upregulated and 1,272 were downregulated (Figure 2C). Functional enrichment analysis indicated that activated various HCC-related pathways, such as the P53 signaling pathway, trypto-phan metabolism, as well as primary bile acid biosynthesis (Figure 2D). The overlapping DEGs may be involved in cellular amino acid catabolic process, carboxylic acid catabolism, and other metabolic processes (Figure 2E).

**Figure 2.**
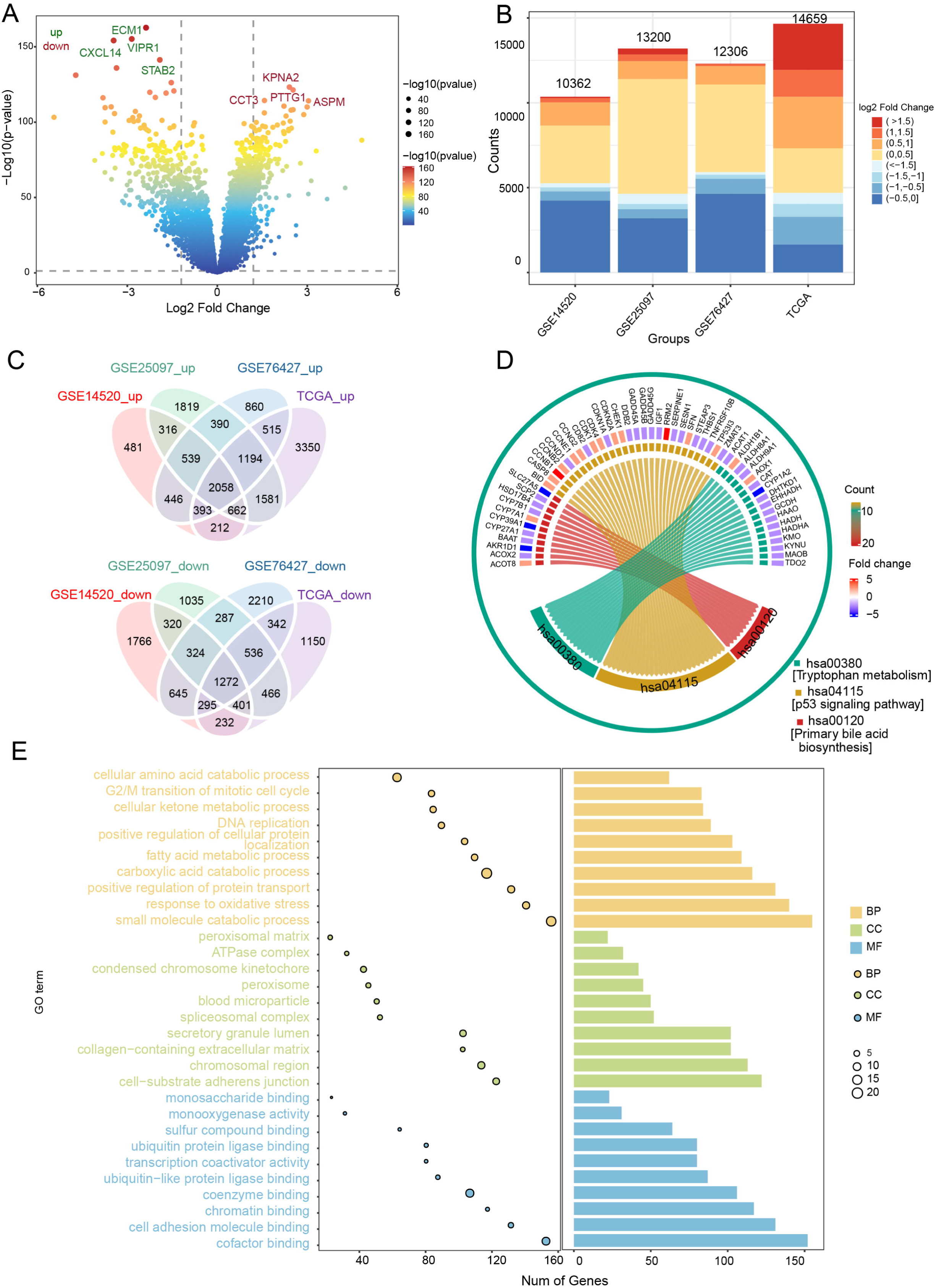
Differentially expressed genes (DEGs) in HCC and their enrichment in biological processes and KEGG pathways. (**A**) DEGs in the GSE14520 dataset. Red indicates upregulated genes and blue, downregulated genes. (**B**) Bar diagram of DEGs in the GSE14520, GSE25097, GSE76427 and TCGA datasets. (**C**) Up- and downregulated DEGs overlapping across the datasets, as visualized in a Venn diagram. (**D**) Overlapping DEGs enriched in various KEGG pathways. (E) GO terms of overlapping DEGs, including BP, CC and MF. The x-axis represents the number of DEGs involved in GO terms, and the y-axis the significantly enriched GO terms. BP, biological process; CC, cellular component; MF, molecular function; GO, Gene Ontology; HCC, hepatocellular carcinoma; KEGG, Kyoto Encyclopedia of Genes and Genomes; TCGA, The Cancer Genome Atlas.

GSEA showed that DEGs positively correlated with the cell cycle, mismatch repair, and DNA replication, but negatively correlated with mineral absorption, PPAR signaling, as well as complement and coagulation cascades (Figure S1A). The AUC for predicting 5-year OS of HCC patients in GSE14520 was 58% (Figure S1B). Cox regression and Kaplan-Meier analyses also showed that 217 of the 3,330 overlapping DEGs were closely associated to HCC prognosis, while further enrichment analysis using Metascape revealed that these prognostic genes were significantly enriched in small molecule catabolism, small molecule biosynthesis, and the mitotic cell cycle (Figure S1C).

### Identification of diagnostic genes in HCC

To evaluate the diagnostic value of prognosis-related genes in TCGA and GSE14520, their AUC values were calculated, and 138 of the 217 prognostic genes with AUC > 0.80 were selected (Figure 3A). Those 138 genes were then subjected to univariate Cox analysis to obtain 10 survival-related genes with relative importance > 0.2 (Figure 3B). Subsequent LASSO regression identified seven feature genes with an AUC of 0.744 for predicting 5-year OS: SORBS2, DHRS1, SLC16A2, RCL1, IGFALS, GNA14, and FANCI (Figures 3C-E). The expression data of the seven feature genes were then integrated into three clusters of 127 combinations using the GMM model, and the cluster with the highest AUC was selected to identify feature genes with strong power for predicting HCC recurrence. The average accuracy of the seven feature genes in one of the 127 combinations was 0.9901, as determined by the GMM classifier (Figure 3F). Further investigation of the independent prognostic value of the feature genes by univariate Cox regression analysis indicated that FANCI was significantly associated with poor OS (hazard ratio > 1) in both datasets (Figures 3G-H), suggesting that it may serve as a novel predictive biomarker of HCC recurrence.

**Figure 3.**
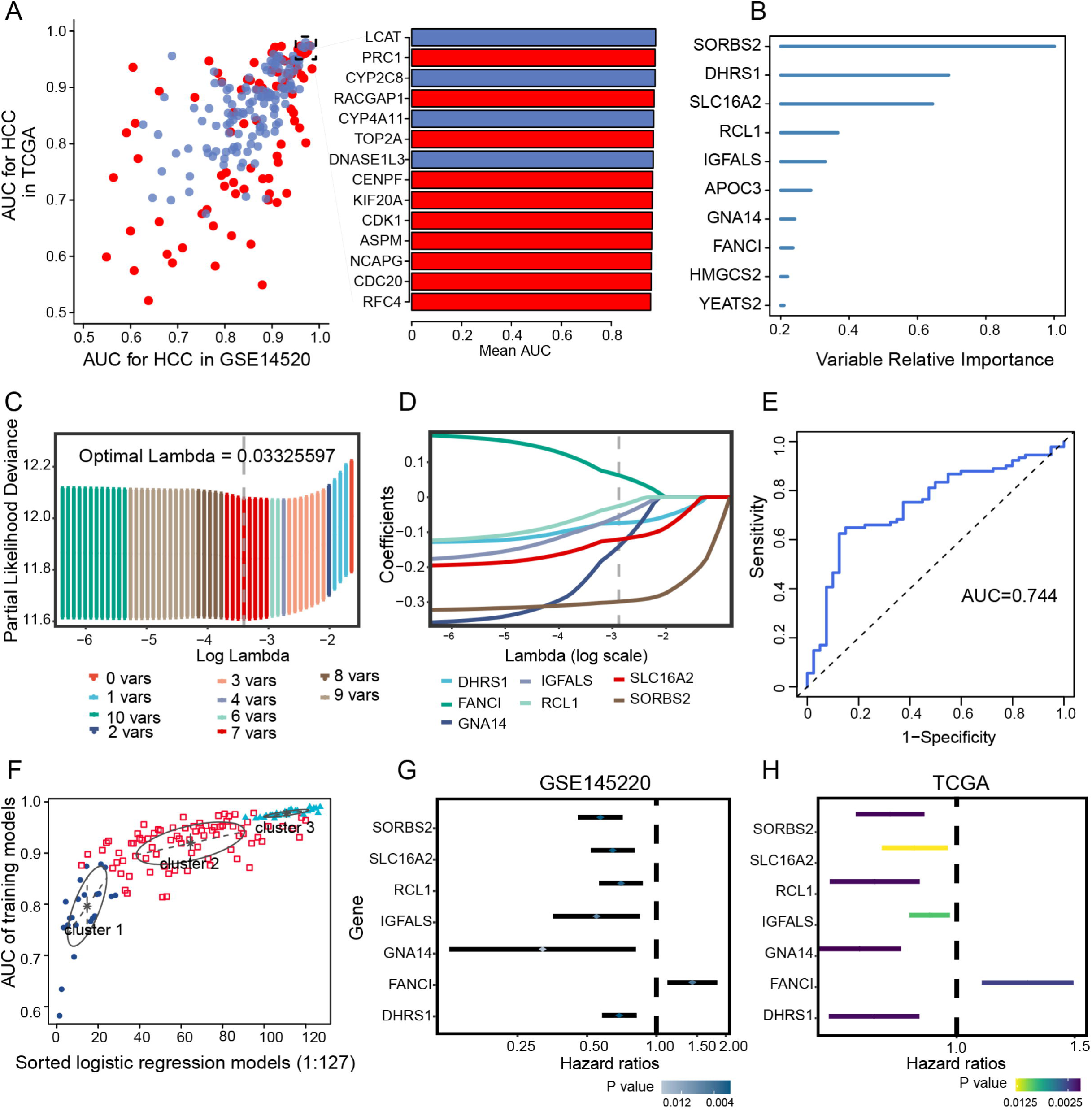
Univariate Cox and LASSO regression. (**A**) Evaluation of the diagnostic value of overlapping differentially expressed genes based on AUC. The 14 genes shown (red, upregulated; blue, downregulated) had AUCs > 0.95. (**B**) Survival-related genes identified by univariate Cox regression analysis of 138 prognostic genes with AUC > 0.80. (**C**) LASSO coefficient profiles of the 10 survival-related genes. (**D**) Ten-fold cross-validation of parameter selection in the LASSO model. (**E**) Time-dependent receiver operating characteristic curves of 5-year overall survival in HCC based on the seven feature genes. (**F**) Pattern of the Gaussian finite mixture model correlated with the AUC scores. There were three clusters of 127 combinations. (**G**,**H**) Univariate analysis of feature genes in the (G) GSE14520 and (H) TCGA datasets. AUC, area under the receiver operating characteristic curve; HCC, hepatocellular carcinoma; LASSO, least absolute shrinkage and selection operator; TCGA, The Cancer Genome Atlas.

### Feature gene-based prognostic risk score as a prognostic tool in HCC

HCC patients were divided into high- and low-risk groups according to the median risk score (Figure 4A-B) and their AUC values for predicting 1-, 3- and 5-year OS were greater than 0.65 for GSE14520 and TCGA data (Figures 4C-D). OS prediction was quantified using nomograms that integrated feature genes with clinicopathological risk factors (Figure 4E). Calibration plots also showed that the nomograms performed well against an ideal model (Figure 4F).

**Figure 4.**
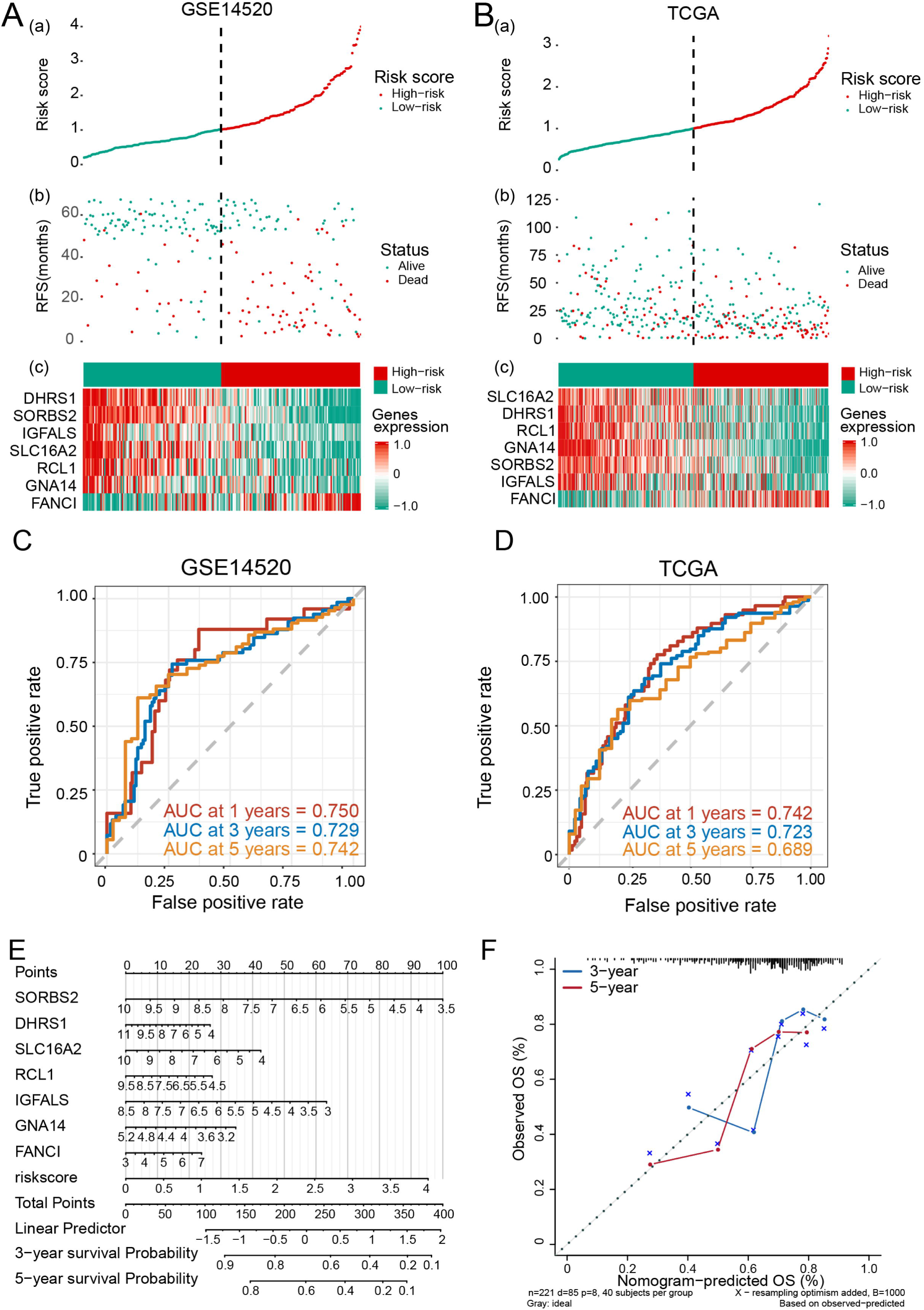
The feature gene-based risk score serves as a prognostic tool in hepatocellular carcinoma (HCC). (**A**,**B**) (a) Risk score, (b) survival status, and (c) expression of the seven feature genes in high-risk and low-risk HCC patients from the GSE14520 and (B) TCGA datasets. (**C**,**D**) Time-dependent receiver operating characteristic curves of 1-, 3-, 5-year overall survival of patients in the (C) GSE14520 and (D) TCGA datasets. (**E**) Quantification of overall survival using nomograms. Lines are drawn upward to determine the points received from the predictor. The sum of these points is reported on the ‘Points’ axis. A line is then drawn downward to determine the 3- and 5-year survival probability based on the seven feature genes. (**F**) Calibration plots showing the performance of nomograms with an ideal model for 3- and 5-year survival. RFS, recurrence-free survival; TCGA, The Cancer Genome Atlas.

### Identification of HCC subtypes by NMF of prognostic genes

In order to identify HCC molecular subtypes, HCC samples from the GSE14520 and TCGA databases were clustered by NMF based on the 217 prognosis-related genes (Figures 5A-C). We were able to divide HCC patients into two molecular subtypes (Figures 5C and S2A): subtype C1 with poor prognosis for HCC, and subtype C2 with good OS (Figures 5E and S2C). We found that the silhouette width value was 0.85 in GSE14520 (Figure 5D) and 0.88 in TCGA (Figure S2B), which suggested good correlation between the HCC samples and the two different subtypes.

**Figure 5.**
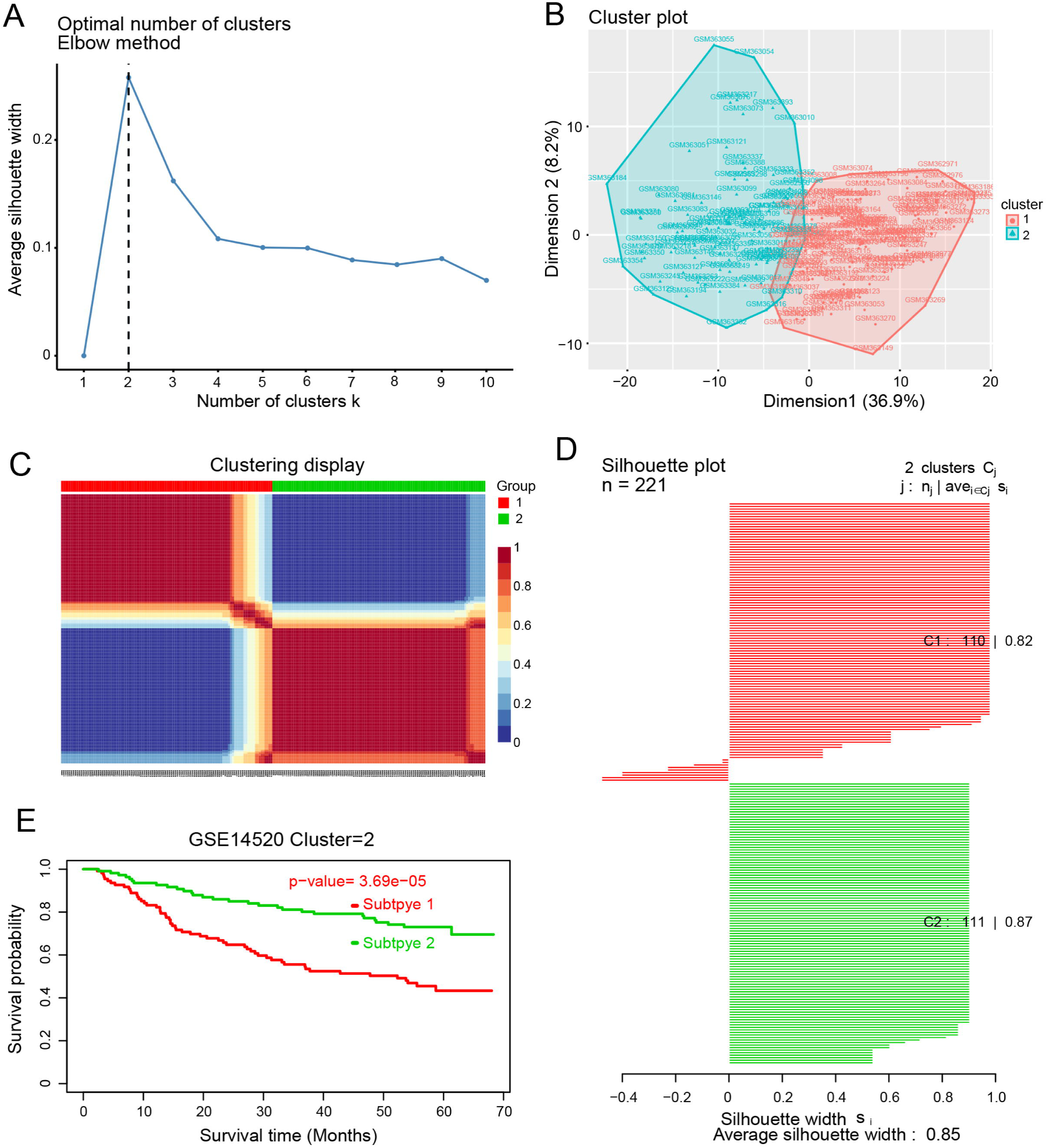
Identification of hepatocellular carcinoma (HCC) subtypes based on the GSE14520 dataset. (**A**) Identification of the optimal number of clusters (k) and visualization of cluster results using the “factoextra” package. (**C**) HCC samples were clustered by non-negative matrix factorization. (**D**) Silhouette width plots. (**E**) Evaluation of the different survival patterns between subtypes using the *CancerSubtypes* package.

### Sensitivity of HCC subtypes to immunotherapy and chemotherapeutic drugs

Comparison of clinical data distribution between the two HCC subtypes in GSE14520 and TCGA indicated that there was no significant difference in age between the two subtypes, but men were more prone to both subtypes of disease than women. In addition, subtype C1 showed shorter survival than subtype C2 (Figure 6A), as well as greater responsiveness to CTLA4-R therapy, based on data from GSE14520 (nominal *P* value = 0.03; Figure 6B) and TCGA (nominal *P* value = 0.09; Figure 6C). In contrast, subtype C2 in TCGA showed significantly greater responsiveness to anticancer drugs ZM.447439 and AG.14699 than subtype C1 (Figure 6D).

**Figure 6.**
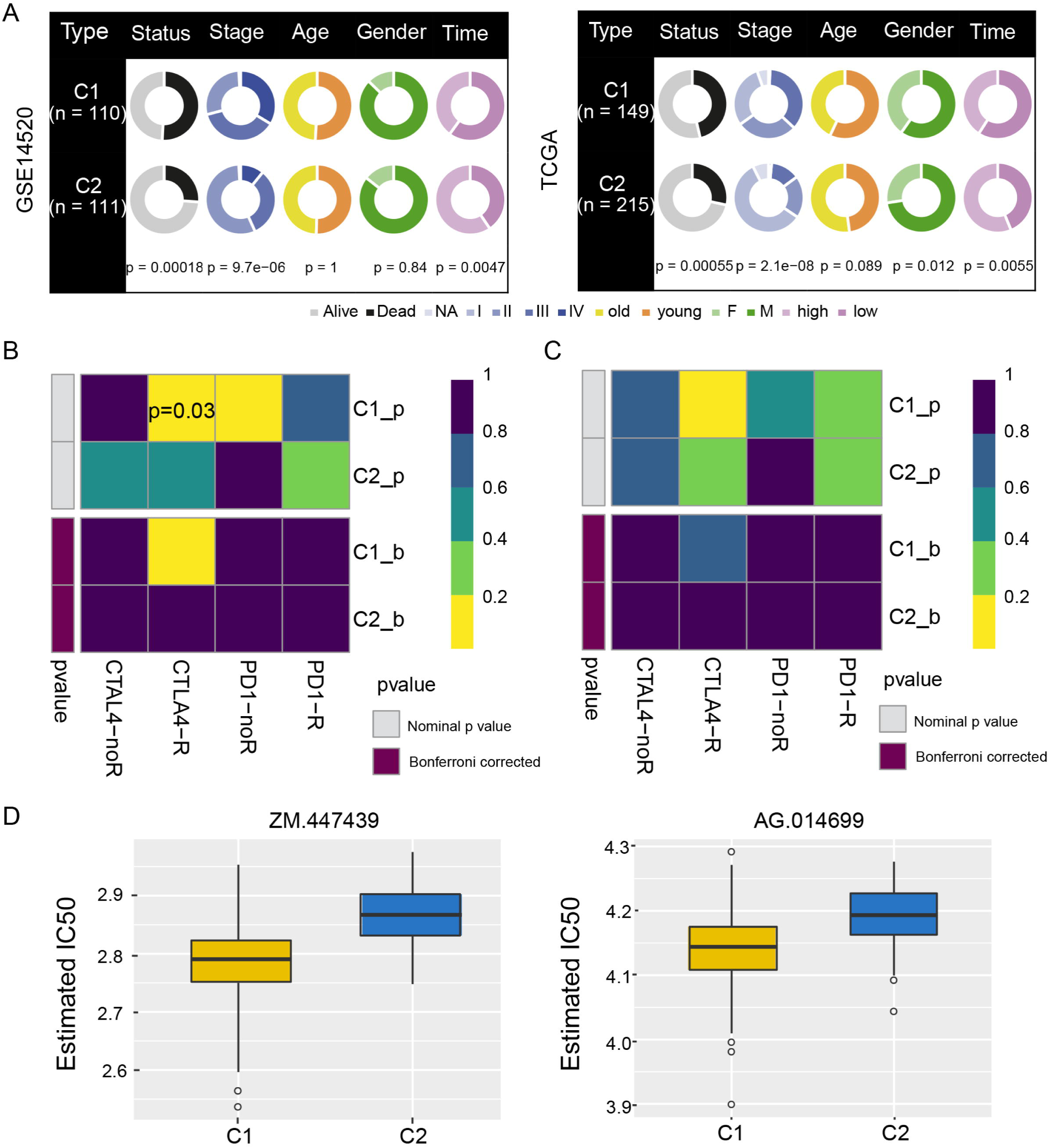
Drug sensitivity and immunotherapy response of hepatocellular carcinoma subtypes C1 and C2. (A) Pie charts comparing the distribution of clinical data between the two subtypes in the GSE14520 and TCGA datasets. (**B**,**C**) Responsiveness of subtypes to immunotherapy in (B) GSE14520 and (C) TCGA, as determined by the Tumor Immune Dysfunction and Exclusion and the SubMap module of the GenePattern database. (**D**) Responsiveness of subtypes in TCGA to anticancer drugs ZM.447439 and AG.014699, as determined by a ridge regression model according to data in the Genomics of Drug Sensitivity in Cancer. TCGA, The Cancer Genome Atlas.

### Stemness index and FANCI expression

Ranking of the HCC samples according to stemness index values showed that their clinico-demographic features significantly correlated with mRNAsi (Figure S3A). FANCI positively correlated with the stemness index (Figure S3B), and it was upregulated in both the GSE138178 and GSE84006 datasets (Figures S3C-D). Further analysis of FANCI mRNA expression in various cancer types using the Oncomine database showed that FANCI was significantly upregulated in liver cancer compared to normal tissues (Figure S3E). These results were confirmed by TCGA RNA-sequencing data in TIMER, which indicated that FANCI levels were significantly higher in tumors than in adjacent normal tissues (Figure S3F).

### Enrichment of FANCI in biological pathways

To explore the role of FANCI in HCC prognosis, we performed time-ROC analysis, which showed that the AUC for predicting 5-year OS was highest in the GSE14520 dataset (Figure 7A), while the AUCs for predicting 1-, 3-, 5-year OS were greater than 0.60 in TCGA (Figure 7B). The AUC for FANCI was higher than 0.90 in all four datasets (Figure 7C). Enrichment analysis showed that FANCI positively correlated with the regulation of fibrinolysis, epoxygenase P450 pathway, and protein activation cascade, while it negatively correlated with mismatch repair, centrosome separation, and translesion synthesis (Figure 7D). In addition, FANCI positively correlated with cell cycle, DNA replication, and the proteasome, while it negatively correlated with FOXO, IL-17, and p53 signaling pathways (Figure 7E). We also found that FANCI expression was higher in HCC tissues than controls in the Human Protein Atlas database (https://www.proteinatlas.org/) (Figure 8A), and that it may positively regulate the Fanconi anemia pathway (Figure 8B).

**Figure 7.**
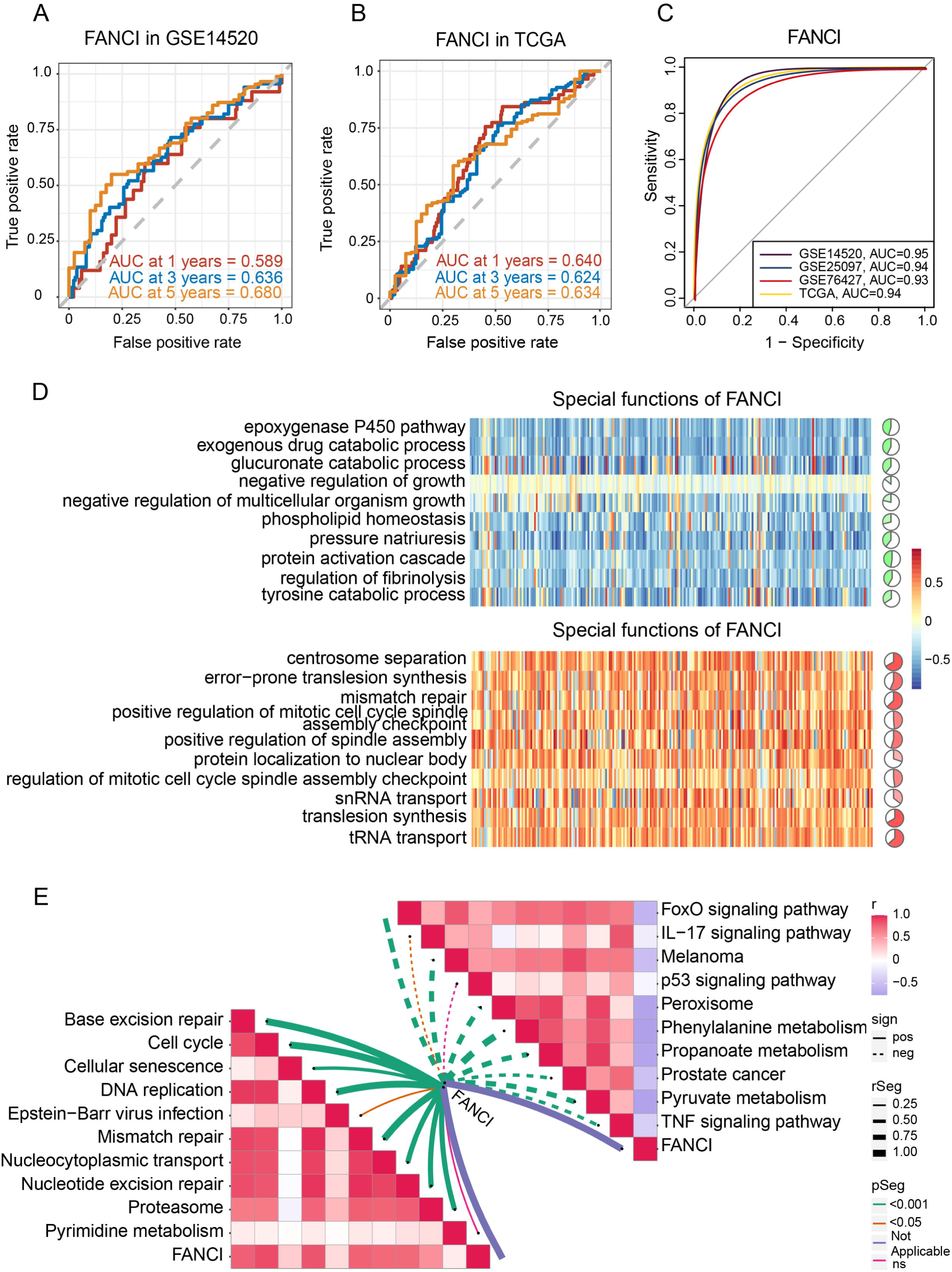
Time-ROC analysis and correlation of FANCI with biological and KEGG pathways. (**A**,**B**) Effect of FANCI on the 1-, 3-, and 5-year overall survival in the (A) GSE14520 and (B) TCGA datasets. (**C**) Effect of FANCI on the AUC values in different datasets. (**D**) Correlation of FANCI with biological processes (red, positive correlation; green, negative correlation). (**E**) Correlation of FANCI with KEGG pathways (red, positive correlation; green, negative correlation). AUC, area under the receiver operating characteristic curve; KEGG, Kyoto Encyclopedia of Genes and Genomes; ROC, receiver operating characteristic curve. TCGA, The Cancer Genome Atlas.

**Figure 8.**
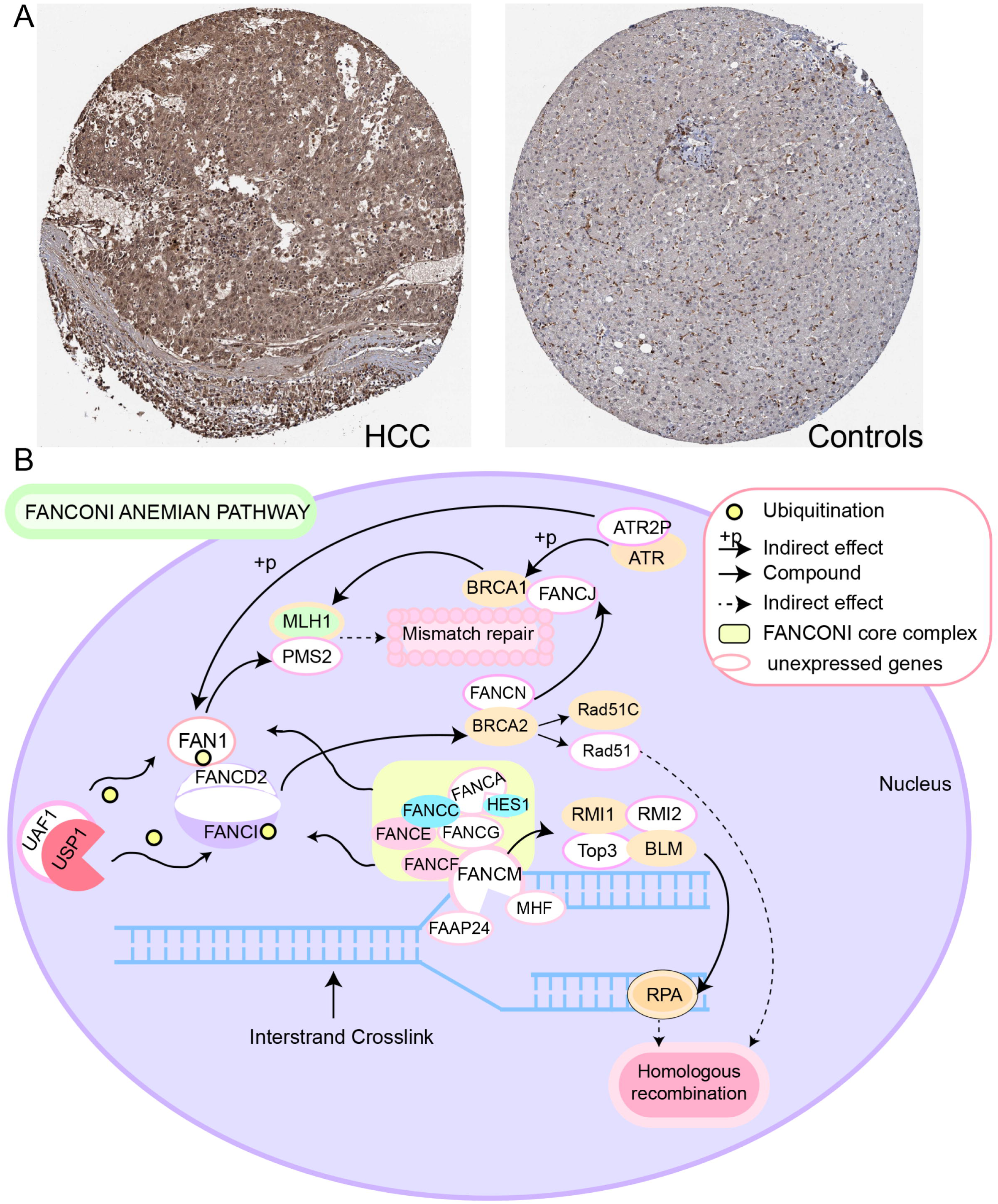
Potential regulatory mechanism of FANCI in hepatocellular carcinoma (HCC). (**A**) Expression of FANCI in HCC and normal tissues obtained from the Human Protein Atlas database. (**B**) Role of FANCI in the Fanconi anemia pathway.

### Immune cell infiltration

To explore the potential clinical significance of immune cell infiltration in HCC, we determined the infiltration levels in all four datasets (Figure S4A). T helper type 2 (Th2), T helper, and pre-dendritic cells were significantly upregulated in HCC samples compared to controls, while T cells and cytotoxic cells correlated significantly with subtype C1 (Figure S4B). Correlations among the 24 immune cell types in HCC tissues were also analyzed (Figure S4C), and four clusters were constructed showing positive and negative correlations with one another (Figure S4D). In addition, dendritic and Th12 cells correlated with the seven feature genes, while a significant correlation was observed between Th2 cells and FANCI (Figure S4E). The CIBERSORT algorithm also showed that most infiltrated immune cells were T cells (Figure S4F).

### Somatic mutations and DNA methylation

Somatic single-nucleotide variants were identified in 364 HCC patients based on sequencing data showing at least 20-fold coverage. Mutations were found to inactivate the tumor suppressor genes *TP53* (31% of all patients), *AXINI* (8%), and *RB1* (5%) (Figure S5A). In addition, we found that FANCI was mutated at two sites in HCC patients, and those mutations may affect protein function (Figure S5B). We further found that FANCI mRNA levels negatively correlated with protein levels, implying that the gene is subject to methylation (Figure S5D). Analysis of genes differentially methylated between HCC and non-tumor tissues in Peruvian hepatocellular carcinoma patients in GSE136319 showed that methylation modification was investigated (Figure S5C). Furthermore, the MEXPRESS tool showed that the methylation level in the promoter region of FANCI was significantly higher in HCC samples than in normal tissues in TCGA (Figure S6).

## Discussion

HCC is one of the most common tumors in the world, but it remains a fatal disease due to its poor prognosis, highlighting the need to identify HCC biomarkers. In this manuscript, we used bioinformatics to determine prognosis-related molecular subtypes of HCC using four public datasets. Our analysis identified two subtypes of HCC (C1 and C2) and seven feature genes that may serve as potential biomarkers and therapeutic targets for HCC. Among them, FANCI showed good prognostic performance and was positively associated with the cell cycle, DNA replication, and mismatch repair.

The identification of novel HCC biomarkers remains critically important. For instance, MITD1 has been reported as a novel liver cancer biomarker involved in cytokinesis [34]. We found that the DEGs overlapping across four databases were involved mainly in the cell cycle, FOXO signaling pathway, mismatch repair, as well as complement and coagulation cascades. Consistent with our results, another study identified DEGs in HCC that were significantly enriched in mismatch repair and complement and coagulation cascades [35]. Juglanthraquinone, a natural compound, can induce apoptosis in HCC cells by activating the Akt/FOXO signaling pathway [36].

In the present study, random forest survival and LASSO regression models identified SORBS2, DHRS1, SLC16A2, RCL1, IGFALS, GNA14, and FANCI as feature genes that may be involved in HCC occurrence and may influence prognosis. Earlier studies showed that IGFALS might be a useful diagnostic and therapeutic target for HCC [37] and that SORBS2 can accurately predict prognosis of HCC patients [38]. IGFALS has also been identified as a tumor suppressor gene, which is silenced by methylation in HCC [39, 40]. For their part, DHRS1 [41], SLC16A2 [42], and GNA14 [43] are significantly underexpressed in HCC tissues compared to normal tissues and have shown potential as prognostic biomarkers of HCC. In addition, RCL1 has shown strong potential for predicting overall and disease-free survival of HCC patients [44], while FANCI has been identified as a reliable marker of hepatitis B virus-associated HCC [45]. Our study showed that SORBS2, FANCI, DHRS1, and IGFALS can be mutated in HCC, and the effects of these mutations should be explored in future work.

Since tumors in different regions may represent different subtypes [10], the identification of HCC subtypes may improve the prognosis of HCC patients and provide new therapeutic strategies. The histological subtypes of primary liver cancer are mainly HCC and intrahepatic cholangiocarcinoma confined to the liver [46]. HCC has recently been stratified into three subtypes differing in metabolic and signaling pathways, including altered kynurenine metabolism, Wnt/β-catenin-associated lipid metabolism, and PI3K/AKT/mTOR signaling [11]. Another study defined three other major HCC subtypes: mitogenic and stem cell-like tumors with chromosomal instability, CTNNB1-mutated tumors displaying immune suppression, and metabolic disease-associated tumors [47]. Clustering of immune cells in the HCC microenvironment led to yet another classification of subtypes as immunocompetent, immunodeficient, or immunosuppressive [12]. In the present work, we identified two molecular subtypes of HCC patients that were associated with different prognosis. Comparison of their clinico-demographic features showed that subtype C1 had significantly shorter OS than subtype C2, consistent with the results of the TCGA groups. In addition, men were more prevalent than women in both subtypes, consistent with a previous study where men showed higher risk of non-alcoholic fatty liver disease and HCC than women [48].

Compared to healthy liver samples, most of the immune cell subpopulations required for antitumor immune response are reduced in HCC samples, whereas gene signatures defining T helper and Th2 cells are significantly increased [49]. In addition, underexpression of tumor antigens in HCC cells reduces T cell activation and tumor infiltration, resulting in a less efficient control of tumor growth, leading to worse clinical outcomes [50]. In the present research, ssGSEA showed that Th2 and T helper cells were significantly upregulated in the four datasets, and cytotoxic and T cells strongly infiltrated tumor tissue in both HCC subtypes.

Our analysis demonstrated that the expression of the seven feature genes positively correlated with dendritic, natural killer, and Th17 cell infiltration. The tumor microenvironment disrupts the maturation and activation of dendritic cells, resulting in dendritic cells with immunosuppressive potential in HCC and breast cancer [51]. Moreover, dysfunction of natural killer cells contributes to HCC development [52], while overexpression of Th17 cells has been associated with worse prognosis of HCC patients [53]. Interestingly, here, we found a significant correlation between FANCI and Th2 cells. Since Th2 cells are associated with immune evasion [54], we hypothesize that FANCI may promote HCC development by evading HCC immune cells.

The seven feature genes are highly expressed in HCC, and their roles should be further studied both *in vitro* and *in vivo*. Further studies are also needed to experimentally validate the effect of FANCI on postoperative recurrence. In fact, all our bioinformatic findings need to be confirmed in preclinical studies and, ultimately, prospective clinical trials.

## Conclusion

We defined two molecular subtypes of HCC that are associated with different prognosis, and we identified FANCI as a good prognostic indicator in HCC.

### Key Points

- Two subtypes of HCC were identified based on tumor and non-tumor data using non-negative matrix decomposition.
- Genes from four HCC datasets were significantly enriched in the cell cycle, FOXO signaling, as well as complement and coagulation cascades.
- FANCI in HCC positively correlated with the cell cycle, DNA replication, and mismatch repair.
- FANCI was able to predict survival of HCC patients, making it a potential prognostic biomarker.

## Data Availability

All data produced in the present study are available upon reasonable request to the authors
All data produced in the present work are contained in the manuscript
All data produced are available online at The Cancer Genome Atlas (TCGA) database (https://portal.gdc.cancer.gov/) and from the Gene Expression Omnibus (GEO) database (http://www.ncbi.nlm.nih.gov/geo/)

http://www.ncbi.nlm.nih.gov/geo/

## Acknowledgments

The authors are grateful to the patients whose publicly available data made this project possible.

## Authors’ Contributions

Jiazhou Ye and Yan Lin contributed equally to this work. Jiazhou Ye and Yan Lin analyzed the data and drafted the manuscript; Yongqiang Li, Xiaoling Luo and Rong Liang designed the research and revised the manuscript; Xing Gao, Lu Lu and Xi Huang carried out the research; Shilin Huang, Tao Bai and Guobin Wu analyzed the data.

## Data Availability Statement

Data are available in a repository and can be accessed using a unique identifier other than a DOI:The data underlying this article are available in [The Cancer Genome Atlas (TCGA) database and Gene Expression Omnibus (GEO) database, and can be accessed with [GSE138178, GSE14520, GSE76427, GSE25097, GSE84006 and GSE136319] in GEO database.

