## Supplementary Figure 1-6 for "Prognosis-related molecular subtypes and immune features associated with hepatocellular carcinoma"

### Supplementary legends

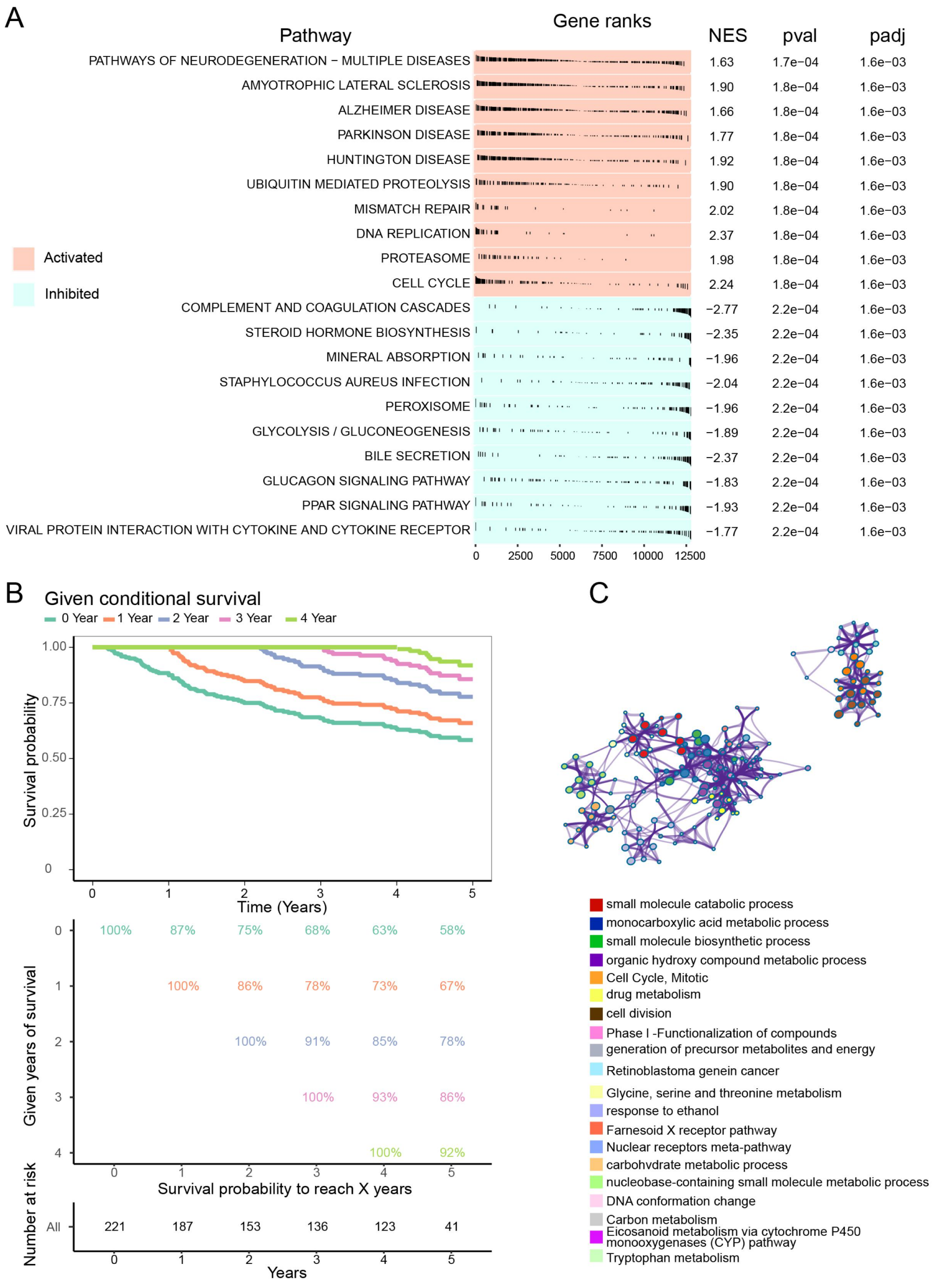


**Figure S1. Gene set enrichment analysis and survival analysis of hepatocellular**

**carcinoma (HCC) samples.** (**A**) Top 10 pathways positively (red) or negatively (blue) correlated with DEGs. (**B**) Kaplan-Meier estimates for conditional survival in HCC patients in GSE14520, assuming a survival rate of 100% at 0–5 years after HCC resection. (**C**) Gene ontology enrichment analysis of the 217 prognostic genes. Each colored dot represents a different biological process.

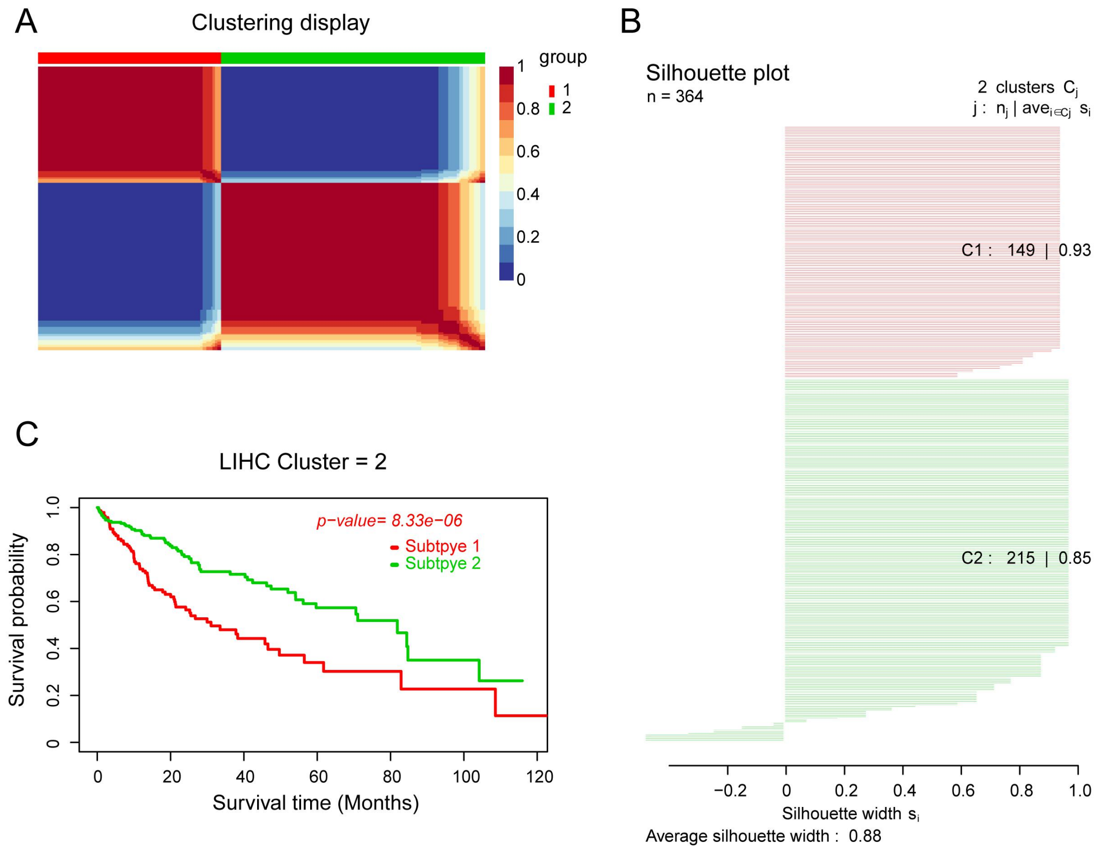


**Figure S2. Identification of hepatocellular carcinoma (HCC) subtypes based on the TCGA dataset.** (**A**) HCC samples were clustered by non-negative matrix factorization. (**B**) Silhouette width plots. (**C**) Evaluation of the different survival patterns between subtypes by the *CancerSubtypes* package. TCGA, The Cancer Genome

Atlas.

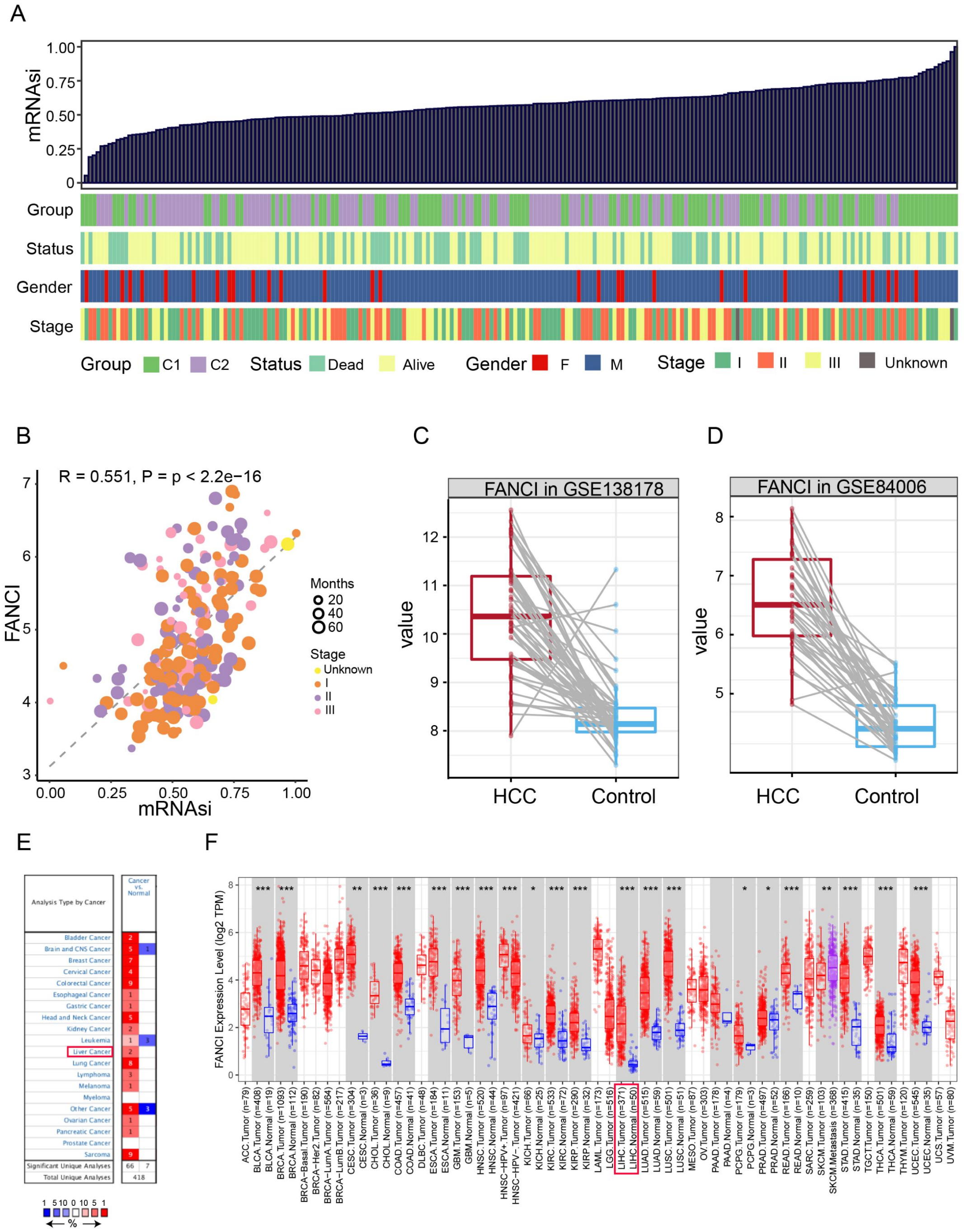


**Figure S3. Clinico-demographic features associated with the mRNA expression-based stemness index (mRNAsi) and FANCI expression in hepatocellular carcinoma (HCC).** (**A**) Correlation of clinico-demographic features (rows) with mRNAsi. Columns in the top row represent samples sorted by mRNAsi from low to high. (**B**) Correlation of mRNAsi and FANCI expression. (**C,D**) FANCI expression in the (C) GSE138178 and (D) GSE84006 datasets. (**E**) FANCI mRNA expression in liver cancer and normal tissues based on the Oncomine database. The number in each cell represents the number of datasets. (**F**) FANCI expression levels in tumor and adjacent normal tissues based on RNA sequencing data from The Cancer Genome Atlas in the Tumor Immune Estimation Resource database. $P < 0.1$, $^{*}P < 0.05$, $^{**}P < 0.01$, $^{***}P < 0.001$.

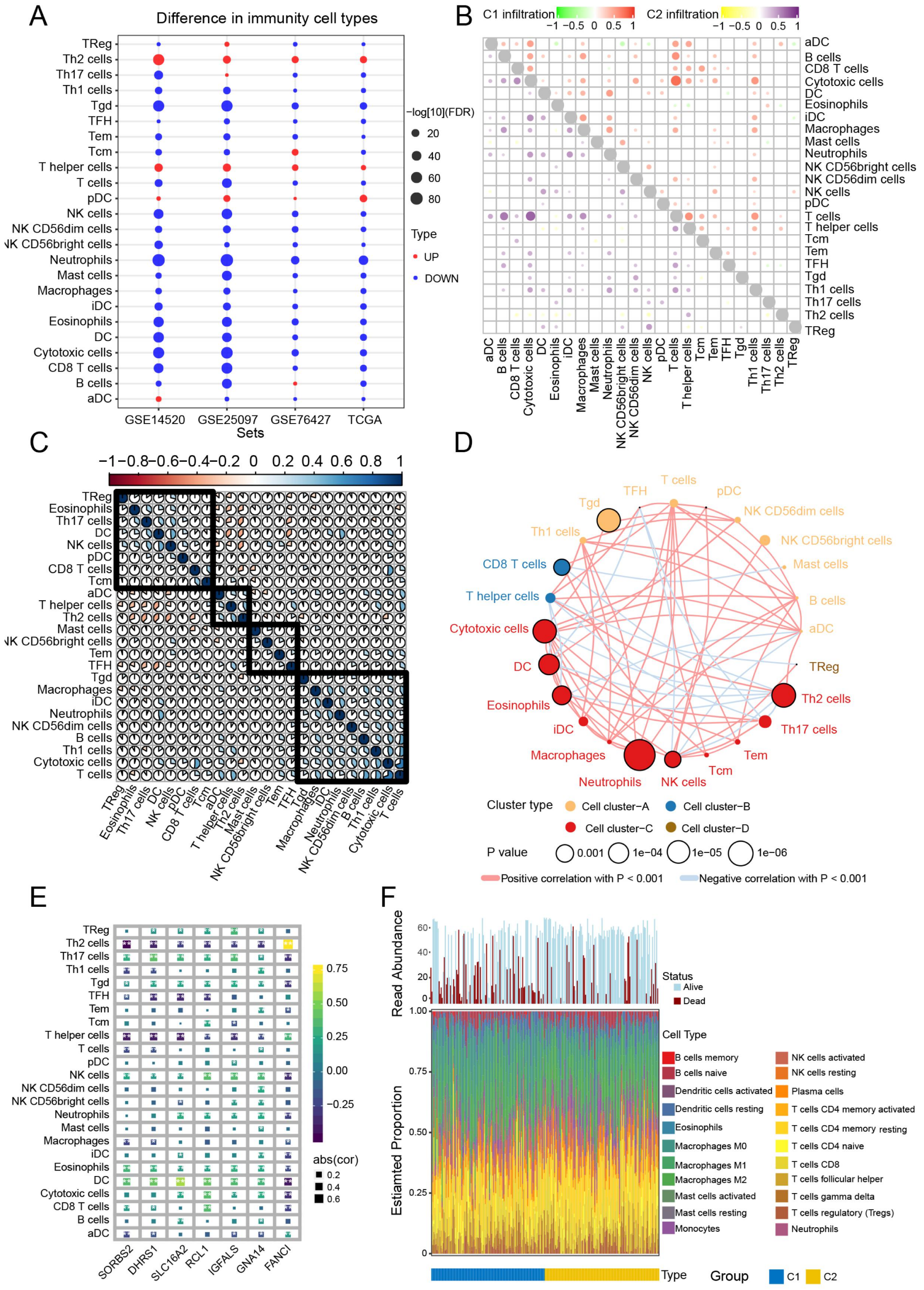


**Figure S4. Infiltration and correlation between immune cells.** (**A**) Levels of immune cell infiltration in the different datasets. (**B**) Correlations among the 24 types of immune cells in the two HCC subtypes C1 and C2. Red and purple represent positive correlations, while green and yellow indicate negative correlations. (**C**) Correlations among the 24 types of immune cells in HCC tissues. Blue sections indicate positive correlation, while orange sections negative correlation. (**D**) Immunity network analysis. Immune cells were grouped into four clusters depending on correlation. Circles represent the prognostic effect of each cell type, and the colour of the line indicates stronger correlation. (**E**) Correlations of immune cell types with the seven feature genes. (**F**) Estimated proportions of 24 immune cell types and survival status in both subtypes. HCC, hepatocellular carcinoma.

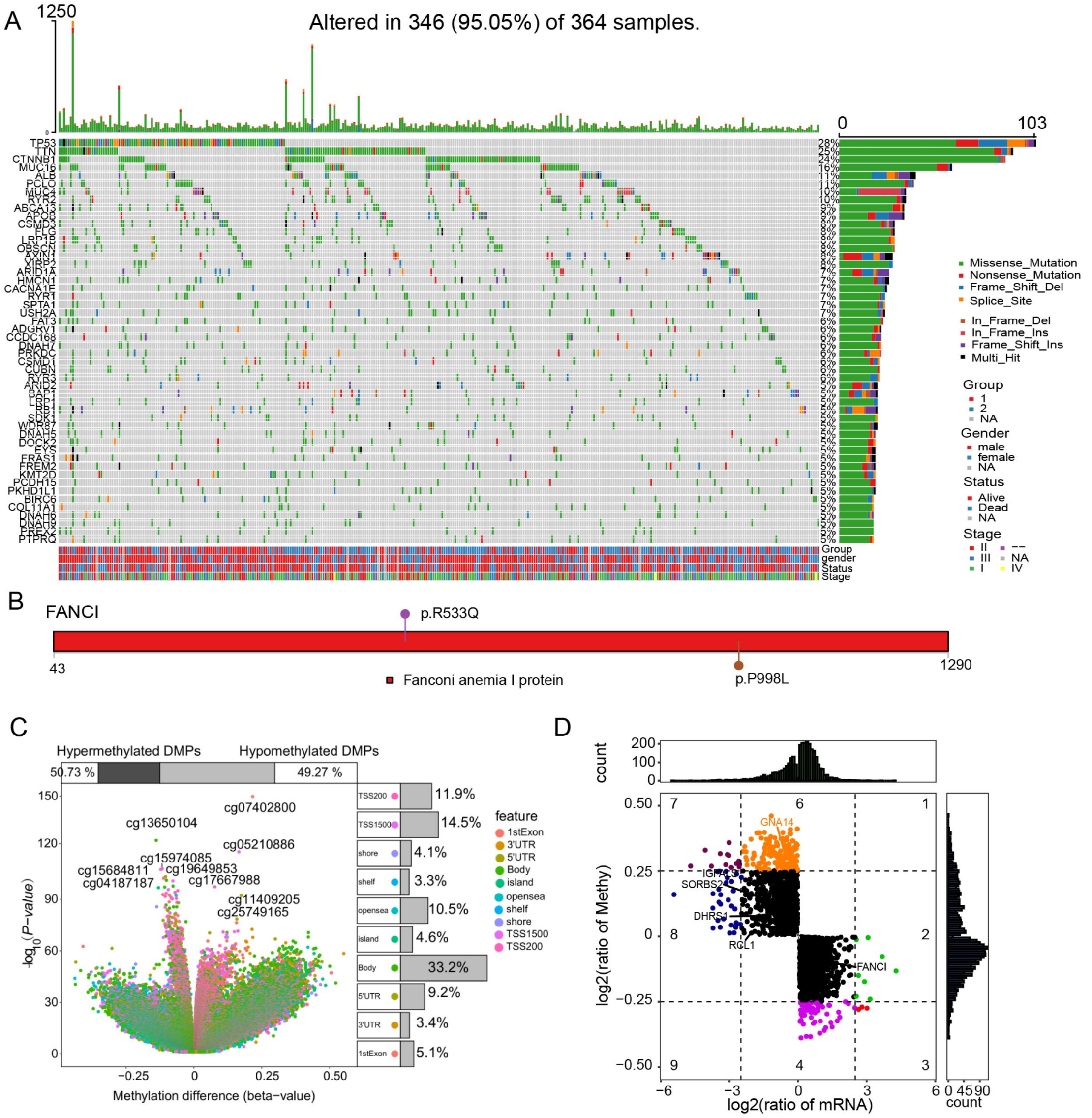


**Figure S5. Genomic landscape of hepatocellular carcinoma and DNA methylation changes.** (**A**) Waterfall plot of somatic mutations in DEGs overlapping across the GSE14520, GSE76427, GSE25097, and TCGA datasets. (**B**) Lollipop graph showing the mutation sites in FANCI in hepatocellular carcinoma. (**C**) Differentially methylated positions (DMPs) of HCC and non-tumor liver tissues obtained from Peruvian hepatocellular carcinoma patients in the GSE136319 dataset. (**D**) Correlation of FANCI mRNA and protein expression. TCGA, The Cancer

Genome Atlas.

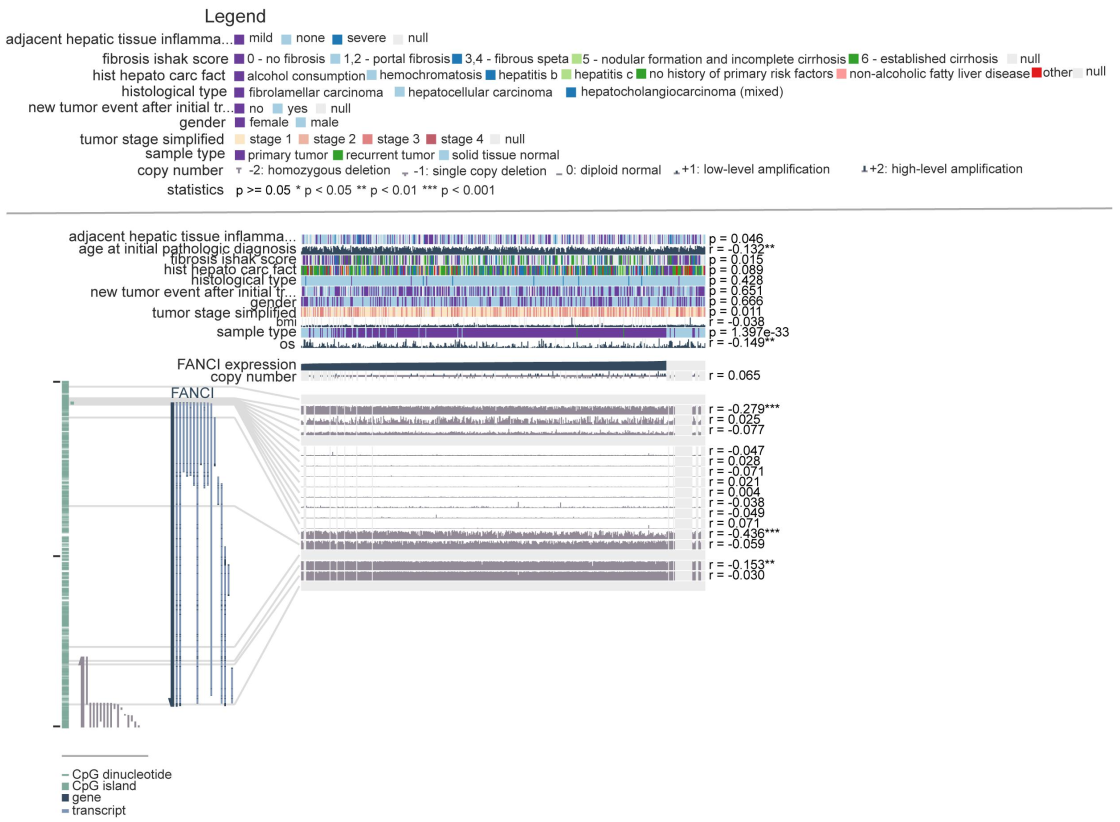


**Figure S6.** Correlation of FANCI expression and methylation status in hepatocellular carcinoma samples from The Cancer Genome Atlas, as determined by the MEXPRESS tool.
